# Low-frequency oscillations in postural sway may have prognostic value for fall risk assessments in aging

**DOI:** 10.64898/2026.02.05.26345669

**Authors:** M. Meyer Vega, HN. Rizeq, DJ. Goble, PE. Gilbert, N. Valadi, N. Baweja, HS. Baweja

## Abstract

The aim of this study was to investigate the effects of cognitive dual-tasking on low-frequency oscillations during quiet standing in older adults. Thirty-two older adults (age 71±8 yrs) were categorized into high- and low-risk fall groups. Both groups performed three trials each of the following tasks: 1) quiet standing with eyes open – on a force plate; 2) quiet standing with eyes open and verbal memory encoding task – on a force plate; and 3) quiet sitting with eyes open and verbal memory encoding task – not on a force plate. We found that: A) older adults at high fall risk exhibit greater postural sway when compared with older adults at low fall risk, B) most of the absolute and normalized wavelet power from 0-4 Hz is concentrated within the 0-1 Hz frequency band across all directions, and C) the absolute change in wavelet power in the 0-1 Hz band from single to dual-task is associated with increased total COP sway displacement irrespective of fall risk group. Based on these findings, it is concluded that nonlinear postural sway measures provide valuable insights into age-associated changes in fall risk and dual-task performance. Focusing on low-frequency oscillations, particularly in the 0-1 Hz band, could enable the earlier identification of individuals at high risk of falls and a better understanding of how the dual-tasking paradigm challenges the aging population.

## Introduction

Postural sway is a proxy for standing balance and a biomarker that identifies the risk of falls [1, 2]. This is particularly useful in older adults, where fall-related injuries represent a significant public health concern [3, 4]. Traditionally, this postural sway is objectively quantified over a period of time (e.g., 20 seconds) by measuring the linear displacement of the center of pressure (COP) signal during quiet standing using force plates or inertial measurement units (IMUs) [1, 2, 5–7]. These devices also measure directional components of the COP sway displacement in the anterior-posterior (AP) and medial-lateral (ML) directions [8, 9]. Alternatively, these signals can be used to investigate the mechanisms of systemic contributions to overall postural control complexity by examining both time and frequency domains [10, 11]. Analysis across these domains allows capturing the multiple timescales that constitute the COP signals [5]. Through this approach, we can identify oscillatory components from multisensory interactions, the effects of external perturbations such as multitasking, and age-associated changes in postural control [11–14].

Postural control primarily relies on the interaction of visual, vestibular, and proprioceptive inputs. The nervous system dynamically adjusts the relative contribution of each sensory input based on environmental conditions through sensory interaction [15, 16]. However, the relative contributions of frequency oscillations constituting postural sway remain unclear. While some groups have identified oscillations in the COP signal as power in frequencies 0-4 Hz [17, 18], others have demonstrated that the most essential information about the underlying mechanisms needed to maintain balance is contained within the 0-1 Hz frequency band [13, 19, 20]. Specifically, distinct frequency bands have been suggested to correspond to different sensory inputs: visual (0-0.1 Hz), vestibular (0.1-0.5 Hz), and proprioceptive (0.5-1 Hz) [13, 19, 21].

Modulation of the power spectrum across these frequency bands can, therefore, provide insights into sensory interactions and information processing. For instance, decreased power in the visual range (0-0.1 Hz) combined with an increase in power in the vestibular and proprioceptive bands (0.1-1 Hz) may indicate a greater reliance on vestibular and proprioceptive inputs relative to visual information, which could also be task dependent [5, 22]. Thus, multispectral analysis of COP time-series signals is valuable in discriminating patterns across different frequency domains, potentially revealing patterns of underlying postural control mechanisms and age-associated changes [11, 23, 24].

Moreover, increased postural sway is linked to a higher risk of falls in older adults [1, 2, 25, 26]. This risk is further exacerbated by age-associated cognitive decline, which directly affects executive cognitive function, working memory, and dual-task performance, all of which contribute to balance disturbances [27, 28]. However, findings on the effect of cognitive dual-tasking on postural control have yielded conflicting results. While some groups have observed decreased postural control during dual-tasking [29, 30], others have demonstrated that postural control remains unperturbed [31]. These inconsistencies may stem from the limitations of traditional linear measures, such as total COP sway displacement and sway area, that quantify the magnitude of COP movement during a specific task [32, 33], but do not provide information about the time-evolving structure of the COP signal [34]. Consequently, nonlinear analysis of the COP signal offers potential value in differentiating patterns of oscillatory responses and sensory interaction associated with fall risk and dual-task performance. Therefore, the purpose of this study was to investigate the effects of cognitive dual-tasking on low-frequency oscillations during quiet standing in older adults.

## Methods

### Participants

Thirty-two older adults (71 ± 8 years, 12 males and 20 females) volunteered to participate in this study (Table 1). All participants completed the Geriatric Depression Scale (GDS) [35]. Individuals with a history of neurological disease, major medical illness (e.g., cancer), head injury with loss of consciousness, current substance abuse, a GDS/BDI-II score indicating moderate or severe depression, or evidence of dementia as revealed through neuropsychological assessment were excluded from this study. The Institutional Review Board at San Diego State University approved the testing procedures, and all participants provided written informed consent before participating in the study.

**Table 1.**
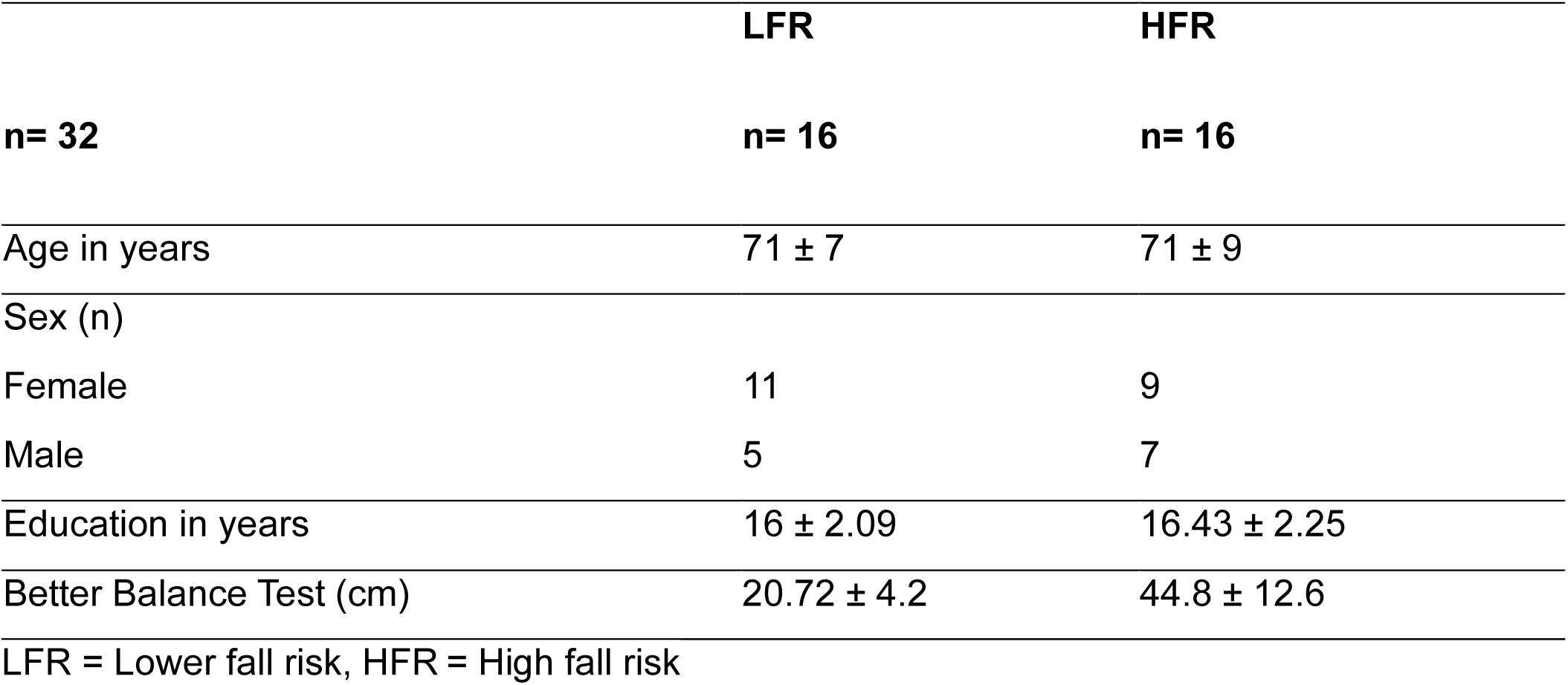
Participants’ demographics, including sample means and standard deviations, are presented.

### Equipment

Postural sway was assessed using the Balance Tracking System BTrackS (Balance Tracking Systems, San Diego, CA, USA). BTrackS consists of a relatively low-cost and portable force plate known as the BTrackS Balance Plate and software (Assess Balance), which has been shown to be a valid and reliable means of acquiring COP data related to postural sway [8, 9, 36, 37]. COP data were collected using the BTrackS in this study with a manufacturer-determined sampling rate of 25 Hz and stored to a local PC for offline analysis.

### Experimental procedures

All participants participated in a single experimental session lasting approximately 1 hour. Each participant began the session with an explanation, demonstration, and familiarization with the testing equipment and protocol. Using the BTrackS Balance and Fall Risk protocol, a measure of participant fall risk was initially made [1, 2]. All participants performed three trials of each of the following tasks: 1) quiet standing with eyes open – on force plate (single-task); 2) quiet standing with eyes open and verbal memory encoding task – on force plate (dual-task); and 3) quiet sitting with eyes open and verbal memory encoding task – off force plate (single-task). Participants were given a three-minute break between each task. All tasks were randomized and blocked across trials (Fig. 1).

**Fig. 1.**
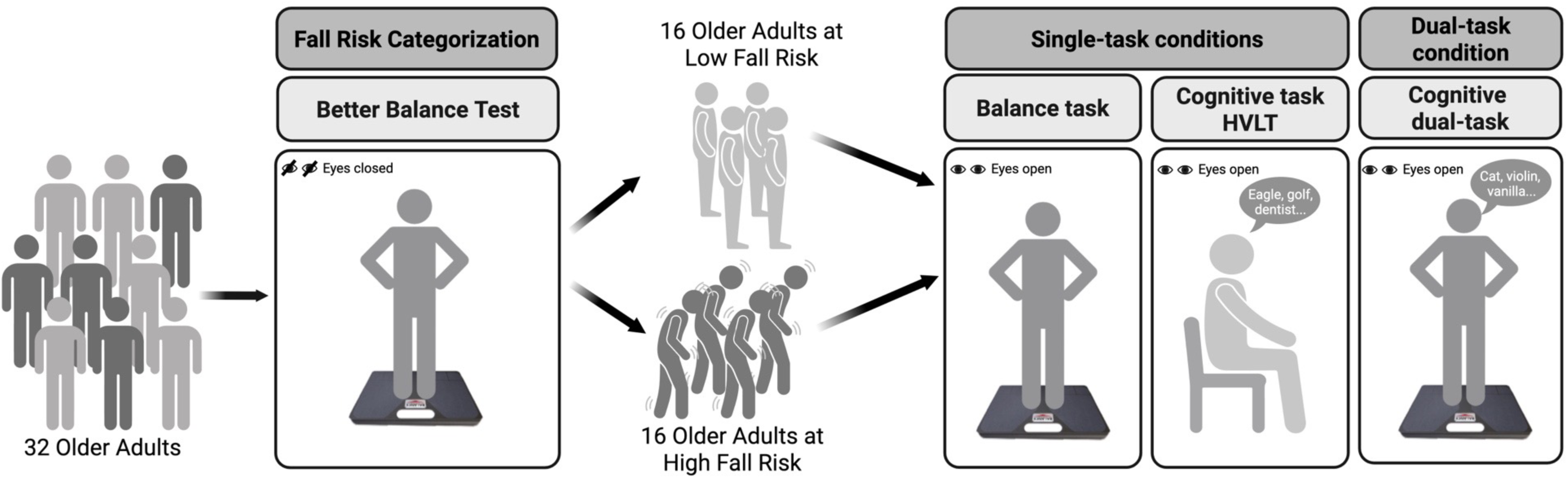
Study design. Thirty-two older adults completed the Better Balance Test for fall risk categorization. Based on their score, participants were classified into low fall risk (n=16) and high fall risk (n=16) groups. All participants completed three experimental conditions: 1) quiet standing with eyes open – on force plate (single-task), 2) quiet standing with eyes open while performing a verbal memory encoding task – on force plate (dual-task condition), and 3) quiet sitting with eyes open while performing the verbal memory encoding task – off force plate (single-task). Schematic created using Biorender.com. HVLT, Hopkins Verbal Learning Test

#### BTrackS Balance and Fall Risk Protocol

For the BTrackS Balance and Fall Risk protocol testing, participants were instructed to stand as still as possible on the force plate across four 20-second trials [1, 2]. The first trial served as a familiarization trial. The succeeding three trials were averaged to calculate the criterion score, which is the total COP sway displacement in cm over 20 seconds, averaged across three trials. Participants with a score of >30 cm or higher were classified as high fall risk (HFR), and those with a score of <30 cm were classified as low fall risk (LFR) (Table 1; Fig. 2). These cut-off scores are based on normative BTrackS data from 16,357 community-dwelling individuals across the United States [1].

**Fig. 2.**
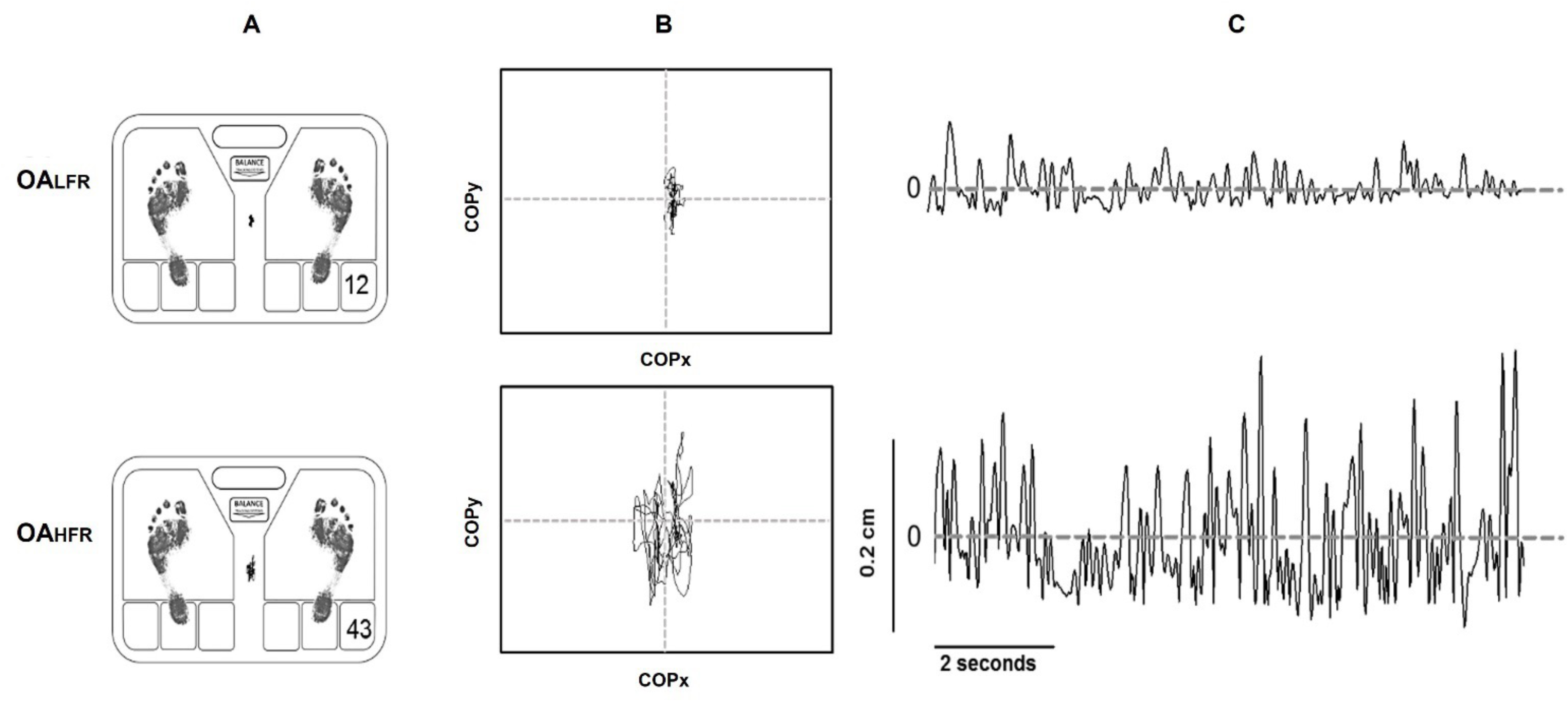
Representative trials under the single-task condition (eyes open). **(A)** Representative trial of older adults at LFR (top panel) and HFR (bottom panel) during the single-task condition. Participants were asked to stand still on the force plate, with their feet shoulder-width apart and their hands on their hips for 20 seconds. **(B)** Raw signal of the total COP sway displacement of older adults at LFR (top panel) and HFR (bottom panel). **(C)** Detrended signal of total COP sway displacement of older adults at LFR (top panel) and HFR (bottom panel)

#### Single and Dual Task Protocols

All participants performed two different single tasks and one dual task in a randomized order.

#### Balance test (Single-task)

Participants stood as still as possible on the BTrackS Balance plate with their hands on their hips, feet shoulder-width apart, and eyes open, looking straight ahead for three 20-second trials.

#### Verbal memory encoding and recall test (Single-task)

Participants performed a modified verbal memory encoding and recall test based on the Hopkins Verbal Learning Test (HVLT) [38]. The first five words from HVLT versions B, D, and F were used during this task for each of the three testing trials. Each trial lasted 20 seconds. The first 10 seconds of each trial consisted of HVLT audio, played back for the participants to memorize in order, followed by 10 seconds of silence. Auditory tones marked the beginning and end of each trial. After each trial, participants were asked to recall the words in the presentation order when balance was not being challenged. Participants were comfortably seated throughout the test. This test consisted of three 20-second trials.

#### Dual-task

This task was designed to assess balance and verbal memory encoding simultaneously. Participants stand quietly on the force plate with their eyes open while undergoing the verbal memory encoding and recall test. For this task, the first five words from the HVLT versions A, C, and E were paired with each standing balance trial on the force plate. After each trial, the participant was asked to recall the words in the correct order when their postural sway was not being measured. We used recall at the end of the trial versus verbalization during the balance trial to negate the effects of vocal articulation on postural sway. Changes in respiration during vocalization have been shown to influence postural sway [39]. This test consisted of three 20-second trials.

### Data analysis

Data were acquired using the BTrackS Assess Balance software and analyzed offline with a custom-written program in MATLAB (MathWorks™, Inc., Natick, MA, USA). The postural sway signal for each 20-second trial length was filtered using a 4th-order Butterworth filter with a low-pass cut-off of 4 Hz. The dependent variables were the anterior-posterior, medial-lateral, and total COP sway displacement (cm). Medial-lateral (ML) and anterior-posterior (AP) sway displacement were calculated using the following formulas [36]:

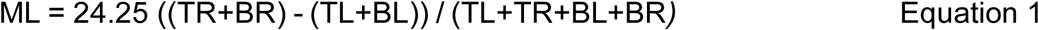

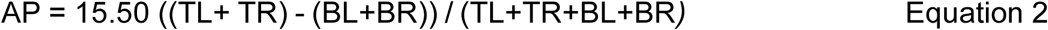

Where TR, TL, BR, and BL are the sensor values from the top right, top left, bottom right, and bottom left corners of the force plate. The constant 24.25 represents half of the ML width of the BTrackS Balance Plate, while 15.50 represents half of the AP length. The total sway displacement is determined by the distance between successive registered COP locations according to the following formula:

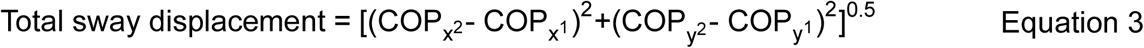

Where COP_x2_ and COP_x1_ are adjacent time points in the COP_x_ (medial-lateral) time-series, and COP_y2_ and COP_y1_ are adjacent time points in the COP_y_ (anterior-posterior) time-series. The total COP sway displacement is then obtained by summing all these individual distances across the entire time series [2].

In addition, continuous wavelet transform analyses were performed on the COP signal using a base MATLAB function developed by Torrence and Compo (available at: http://paos.colorado.edu/research/wavelets) [40]. As part of this analysis, the total COP signal was detrended using MATLAB’s detrend function. The wavelet transform of the COP signal determines both the amplitude-frequency characteristics of the signal and the way its amplitude varies with time [41]. The wavelet represents a set of functions in the form of small waves created by dilations and translations from a simple generator function, Ψ(t), which is called the mother wavelet [42]. To perform a wavelet transform, the Morlet mother wavelet was calculated according to the following formula [40]:

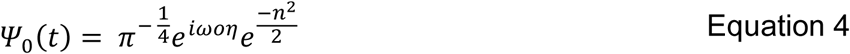

Where η dimensionless time and 𝑤_0_ dimensionless frequency (in this study, we used 𝑤_0_ = 6, as suggested by Grinsted and colleagues [43]. The Morlet wavelet (with 𝑤_0_ = 6) is appropriate when performing wavelet analysis, since it provides a balance between time and frequency localization [43]. The wavelet transform applies the wavelet function as a band-pass filter to the time series. To modify its frequency content, the wavelet function is stretched in time by varying its scale(s) (dilation). For the Morlet wavelet used in this study, the wavelet scale is almost equal to the Fourier period (Fourier period = 1.03 s) and was calculated according to the following formula:

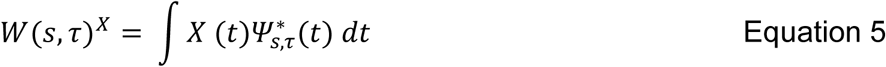

Where 𝑠 represents the dilation parameter (scale shifting), 𝜏 represents the location parameter (time shifting), and the basic function Ψ_𝑠,𝜏_ (*t*) is obtained by dilating and translating the mother wavelet Ψ_0_ (*t*) [42]. For statistical comparisons, the frequency data of the COP signal were divided first into 0-1, 1-2, and 2-4 Hz frequency bands, and then into 0-0.1, 0.1-0.5, and 0.5-1 Hz frequency bands. The dependent variable for the spectral analysis of the COP signal was the absolute peak and normalized wavelet power in the above bands (0-1, 1-2, and 2-4 Hz, and 0-1, 0.1-0.5, 0.5-1 Hz).

### Statistical analysis

A two-way mixed ANOVA model (2 groups: HFR and LFR x 2 tasks: single and dual-task) with repeated measures on all factors compared the differences in means from AP, ML, total COP sway displacement, and 95^th^ percentile COP ellipse area for the two groups between tasks. A three-way mixed ANOVA model (2 groups: HFR and LFR x 2 tasks: single and dual-task x 3 frequency bands: 0-1, 1-2, 2-4 Hz) with repeated measures on all factors compared the absolute and normalized wavelet power spectrum from 0-4 Hz. A similar model (2 groups: HFR and LFR x 2 tasks: single and dual-task x 3 frequency bands: 0-0.1, 0.1-0.5, 0.5-1 Hz) compared the absolute and normalized wavelet power from 0 to 1 Hz. Furthermore, a two-way ANOVA (2 groups: HFR and LFR x 2 tasks: on the force plate and off the force plate) with repeated measures on the tasks compared the differences in cognitive task accuracy when the verbal memory task was administered while standing (on the force plate) and sitting (off the force plate).

Multiple linear regression analyses (stepwise) were used to determine the association between absolute wavelet power in the 0-1 Hz band and total COP sway displacement. And to determine the association between the absolute change in wavelet power in the 0-1 Hz band from single to dual-task to the change in total COP sway displacement. This change was calculated according to the following formula:

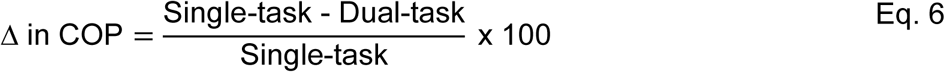

Any participant who exhibited values outside ±3SD was excluded as an outlier (all participants in this manuscript exhibited values within ±3SD). The relative importance of the predictors was estimated with the part correlations (part *r*), which provide the correlation between a predictor and the criterion. The unique contribution of the change in wavelet power from 0-1 Hz to the change in total COP sway displacement from single to dual-task was used to examine the relationship.

All analyses were performed with the IBM SPSS Software (IBM SPSS version 29, SPSS Inc., Chicago, IL, USA), and graphs were created with the Sigma Plot^®^ Software (Inpixon Inc. version 15 for Windows). Appropriate post-hoc analyses were conducted to follow up on significant interactions from the ANOVA models. For example, group-associated differences were followed by independent-sample t-tests. Multiple t-test comparisons were corrected using the Bonferroni corrections, and the alpha level for all statistical tests was 0.05. Data are reported as mean ± SD in text and mean ± standard error of the mean (SEM) in the figures. Only significant main effects and interactions are presented unless otherwise noted.

## Results

### Postural sway

The HFR group exhibited greater AP sway displacement (HFR group 32.84 ± 0.06 cm, LFR group 25.4 ± 0.6 cm; *F*_1,30_ = 26.2, *P* < 0.001, *η_p_*^2^ = 0.467; Fig. 3A), ML sway displacement (HFR group 23.27 ± 0.10 cm, LFR group 21.63 ± 0.12 cm; *F*_1,30_ = 12.1, *P* = 0.002, *η_p_*^2^ = 0.288; Fig. 3B), total COP sway displacement (HFR group 26.52 ± 0.21 cm, LFR group 15.8 ± 0.95 cm; *F*_1,30_ = 25.7, *P* < 0.001, *η_p_*^2^= 0.461; Fig. 3C), and greater COP 95% ellipse area (HFR group 1.53 ± 9.5 cm^2^, LFR group 0.82 ± 0.18 cm^2^; *F*_1,30_ = 8.89, *P* = 0.006, *η_p_*^2^= 0.229; Fig. 3D) when compared with LFR group irrespective of task conditions. All other main effects and interactions were not significant.

**Fig. 3.**
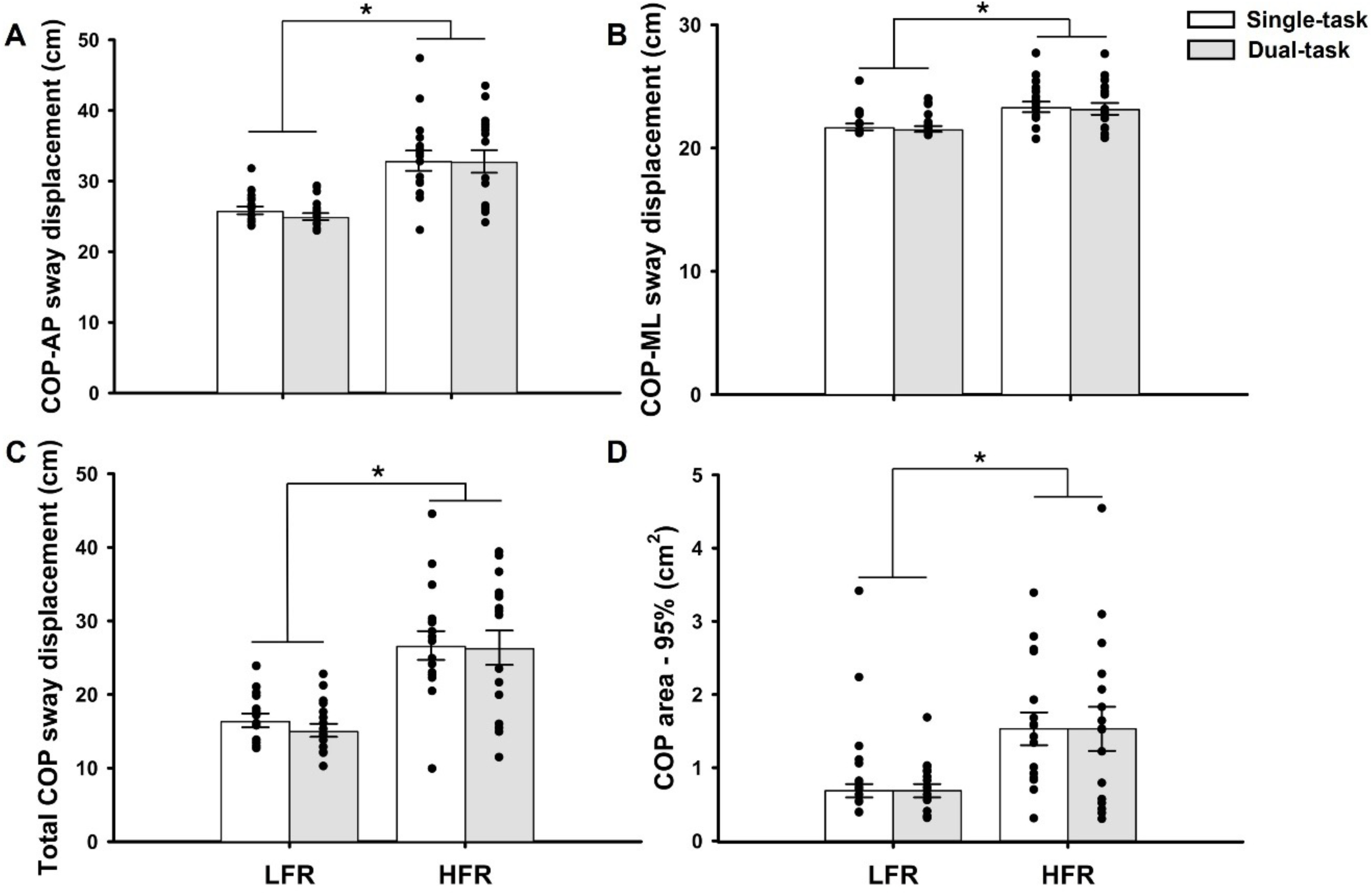
Average postural sway metrics during single and dual-task conditions. **(A)** Older adults at HFR exhibited significantly greater AP sway displacement when compared with older adults at LFR. **(B)** Older adults at HFR exhibit significantly greater ML sway displacement when compared with older adults at LFR. **(C)** Older adults at HFR exhibited significantly greater total COP sway displacement when compared with older adults at LFR. **(D)** Older adults at HFR exhibited significantly greater 95% COP ellipse area when compared with older adults at LFR. * Indicates a significant difference between older adults at HFR and LFR

### Cognitive task accuracy

There was no significant main effect or interaction between the HVLT verbal memory task performance while sitting (HFR group: 3.77 ± 1.01, LFR group: 3.85 ± 1.11, *F*_1,30_ = 0.49, *P* = 0.826, *η_p_*^2^= 0.002) when compared with standing on the force plate (HFR group: 3.42 ± 1.06, LFR group: 3.77 ± 0.93, *P* = 0.323, *F*_1,30_ = 1.01, *P* = 0.323, *η_p_*^2^= 0.033; Fig. 4).

**Fig. 4.**
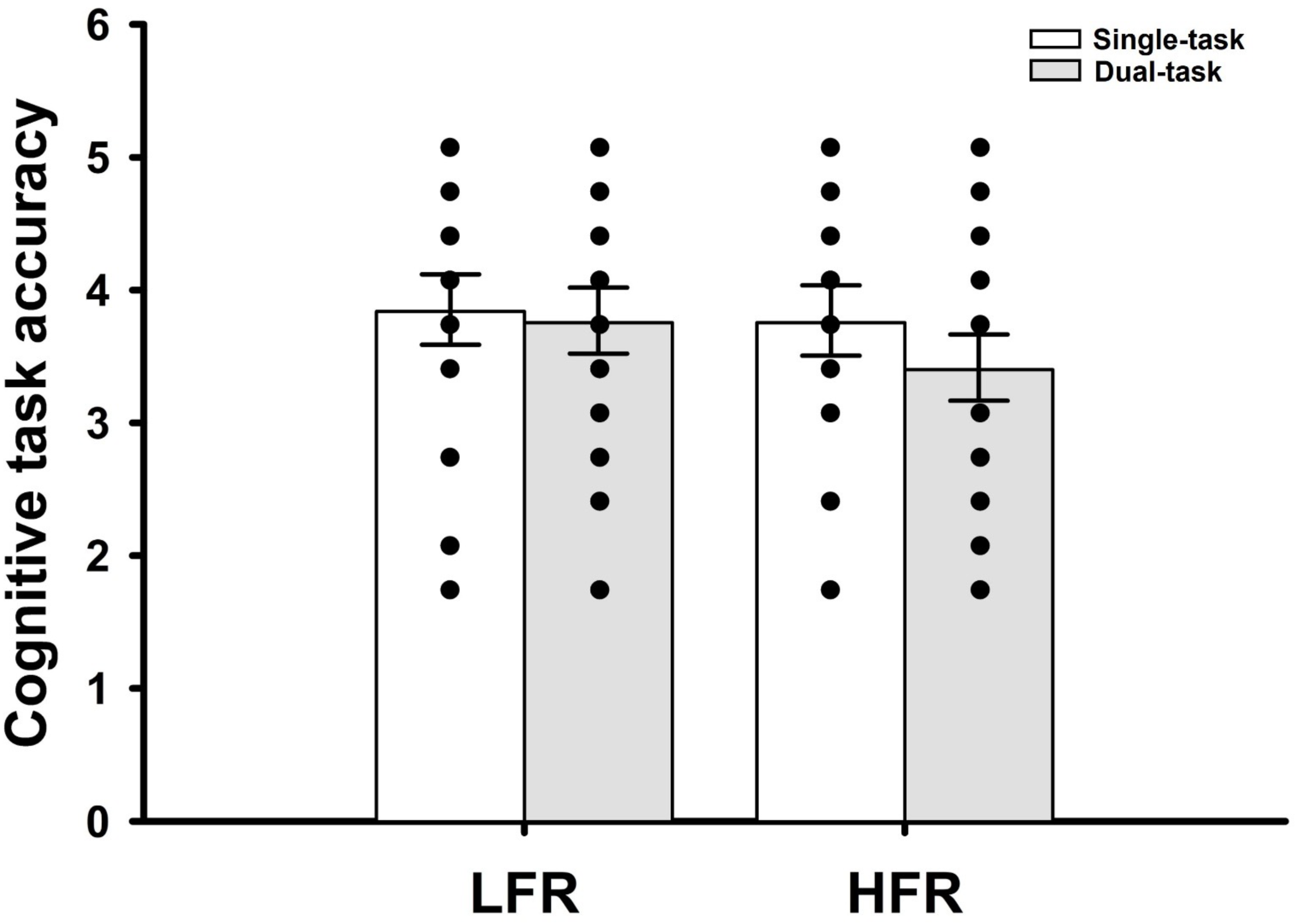
Cognitive task accuracy. There was no significant main effect or interaction between the HVLT verbal memory task while sitting (single-task) when compared with standing on the force plate (dual-task)

Wavelet power spectrum of the center of pressure signal from 0-4 Hz

#### Anterior-posterior 0-4 Hz: absolute and normalized wavelet power

In the absolute wavelet power for AP signal, the frequency main effect was significant (0-1 Hz: 11.858 ± 6.26 cm^2^, 1-2 Hz: 0.290 ± 0.25 cm^2^, 2-4 Hz: 0.087 ± 0.068 cm^2^; *F*_2,60_ = 114, *P* < 0.001, *η_p_*^2^= 0.792; Fig. 5), indicating greater power from 0-1 Hz when compared with 1-2 and 2-4 Hz. In the normalized wavelet power spectrum, the frequency main effect was significant (0-1 Hz: 95.98 ± 3.01 %, 1-2 Hz: 3.03 ± 2.36 %, 2-4 Hz: 0.98 ± 0.84 %; *F*_2,60_ = 12275.54, *P* < 0.001, *η_p_*^2^= 0.998), indicating greater power from 0-1 Hz when compared with 1-2 and 2-4 Hz. All other main effects and interactions were not significant.

**Fig. 5.**
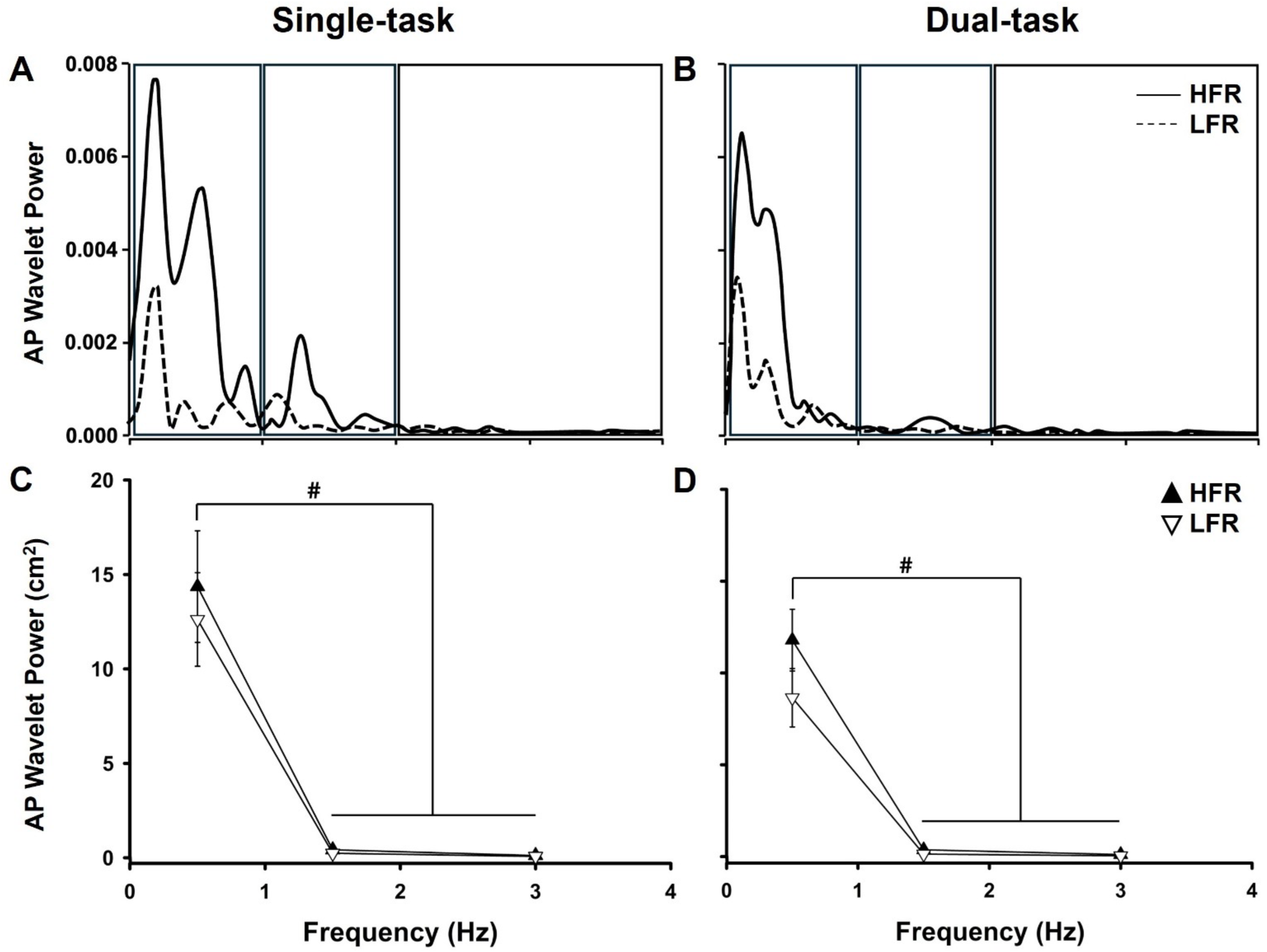
Absolute wavelet power spectrum of AP 0-4 Hz signal. **(A-B)** Representative trial of older adults at LFR (dotted line) and HFR (straight line) during single (left)- and dual-task (right) conditions. **(C-D)** AP wavelet power was significantly greater from 0-1 Hz when compared with 1-2 and 2-4 Hz, irrespective of task conditions. # Indicates a significant difference between frequency bands

#### Medial-lateral 0-4 Hz: absolute and normalized wavelet power

In the absolute wavelet power for ML signal, the frequency main effect was significant (0-1 Hz: 3.34 ± 5.02 cm^2^, 1-2 Hz: 0.067 ± 0.056 cm^2^, 2-4 Hz: 0.016 ± 0.017 cm^2^; *F*_2,60_ = 14, *P* < 0.001, *η_p_*^2^= 0.317; Fig. 6), indicating greater power from 0-1 Hz when compared with 1-2 and 2-4 Hz. In the normalized wavelet power spectrum, the frequency main effect was significant (0-1 Hz: 95.57 ± 2.67 %, 1-2 Hz: 3.72 ± 2.53 %, 2-4 Hz: 0.70 ± 0.26 %; *F*_2,60_ = 13636, *P* < 0.001, *η_p_*^2^= 0.998), indicating greater power from 0-1 Hz when compared with 1-2 and 2-4 Hz. All other main effects and interactions were not significant.

**Fig. 6.**
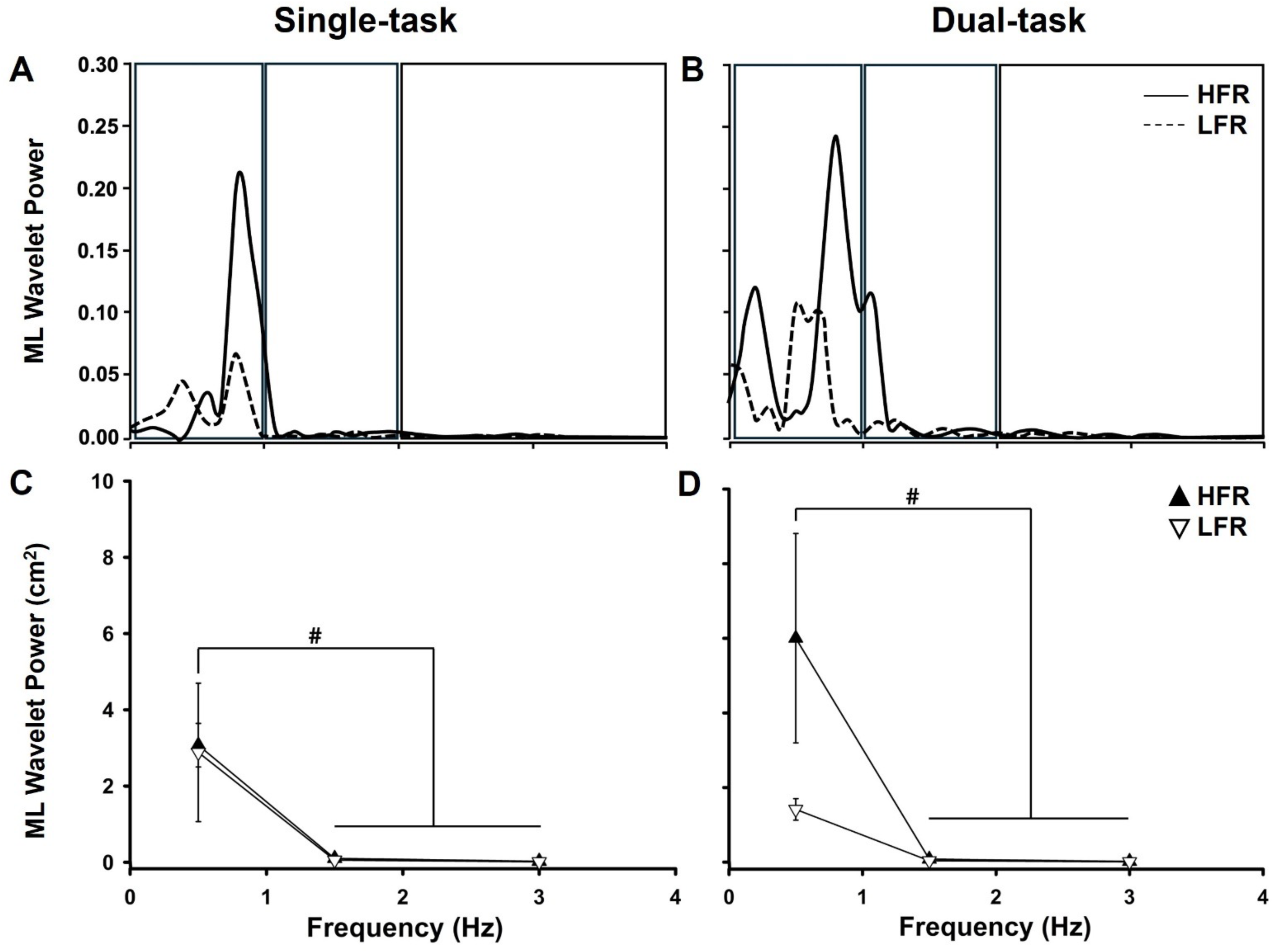
Absolute wavelet power spectrum of ML 0-4 Hz signal. **(A-B)** Representative trial of older adults at LFR (dotted line) and HFR (straight line) during single (left)- and dual-task (right) conditions. **(C-D)** ML wavelet power was significantly greater from 0-1 Hz when compared with 1-2 and 2-4 Hz, irrespective of task conditions. # Indicates a significant difference between frequency bands

#### Total COP 0-4 Hz: absolute and normalized wavelet power

In the absolute wavelet power for the total COP signal, the frequency main effect was significant (0-1 Hz: 0.012 ± 0.006 cm^2^, 1-2 Hz: 0.003 ± 0.0 cm^2^, 2-4 Hz: 0.002 ± 0.0 cm^2^; *F*_1,30_ = 71.6, *P* < 0.001, *η_p_*^2^= 0.705; Fig. 7), indicating greater power from 0-1 Hz when compared with 1-2 and 2-4 Hz. The frequency × group interaction was significant (*F*_2,60_ = 15.1, *P* < 0.001, *η_p_*^2^= 0.335), indicating that the HFR group exhibited greater power across all frequency bands when compared with the LFR group, irrespective of task conditions.

**Fig. 7.**
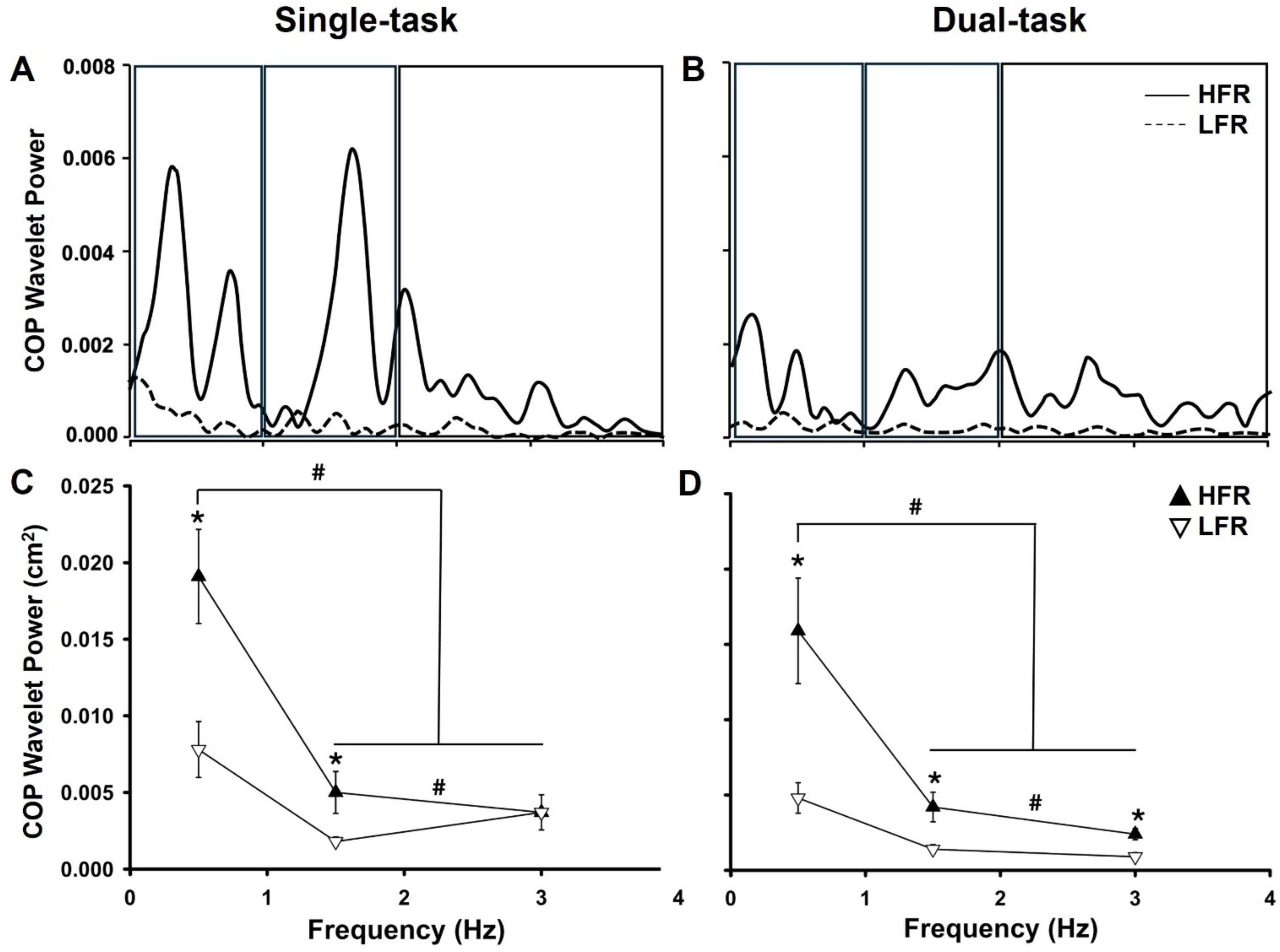
Absolute wavelet power spectrum of the total COP 0-4 Hz signal. **(A-B)** Representative trial of older adults at LFR (dotted line) and HFR (straight line) during single (left)- and dual-task (right) conditions. **(C-D)** Total COP wavelet power was significantly greater from 0-1 Hz when compared with 1-2 and 2-4 Hz, irrespective of task conditions. Older adults at HFR exhibited significantly greater total COP power when compared with LFR. # indicates a significant difference between frequency bands. * Indicates a significant difference between older adults at HFR and LFR

In the normalized wavelet power, the frequency main effect was significant (0-1 Hz: 69.30 ± 9.28 %, 1-2 Hz: 18.94 ± 5.50 %, 2-4 Hz: 11.75 ± 4.81 %; *F*_2,60_ = 451, *P* < 0.001, *η_p_*^2^= 0.938), indicating greater power from 0-1 Hz when compared with 1-2 and 2-4 Hz. All other main effects and interactions were not significant.

#### Wavelet power spectrum of the center of pressure signal from 0-1 Hz

We found that most of the normalized wavelet power of the COP signal from 0 to 4 Hz is concentrated within the 0 to 1 Hz frequency band across all directions. Specifically, ∼96% of the power in anterior-posterior sway, ∼95% in medial-lateral sway, and ∼69% of the power in total COP sway come from the 0-1 Hz frequency range. Therefore, to investigate this further, we analyzed the COP signal within the 0-1 Hz frequency band to examine sensory interaction.

#### Anterior-posterior 0-1 Hz: absolute and normalized wavelet power

In the absolute wavelet power for AP signal, the frequency main effect was significant (0-0.1 Hz: 11.64 ± 6.27 cm^2^, 0.1-0.5 Hz: 6.57 ± 4.58 cm^2^, 0.5-1 Hz: 2.59 ± 2.00 cm^2^; *F*_2,60_ = 52.9, *P* < 0.001, *η_p_*^2^= 0.638; Fig. 8A-B), indicating greater power from 0-0.1 Hz when compared with 0.1-0.5 and 0.5-1 Hz. Task main effect approached significance (*F*_1,30_ = 3.2, *P* = 0.081, *η_p_*^2^= 0.098). In the normalized wavelet power spectrum, the frequency main effect was significant (0-0.1 Hz: 54.81 ± 12.62 %, 0.1-0.5 Hz: 31.89 ± 8.22 %, 0.5-1 Hz: 13.30 ± 7.17 %; *F*_2,60_ = 99.4, *P* < 0.001, *η_p_*^2^= 0.768), indicating greater power from 0-0.1 Hz when compared with 0.1-0.5 and 0.5-1 Hz. All other main effects and interactions were not significant.

**Fig. 8.**
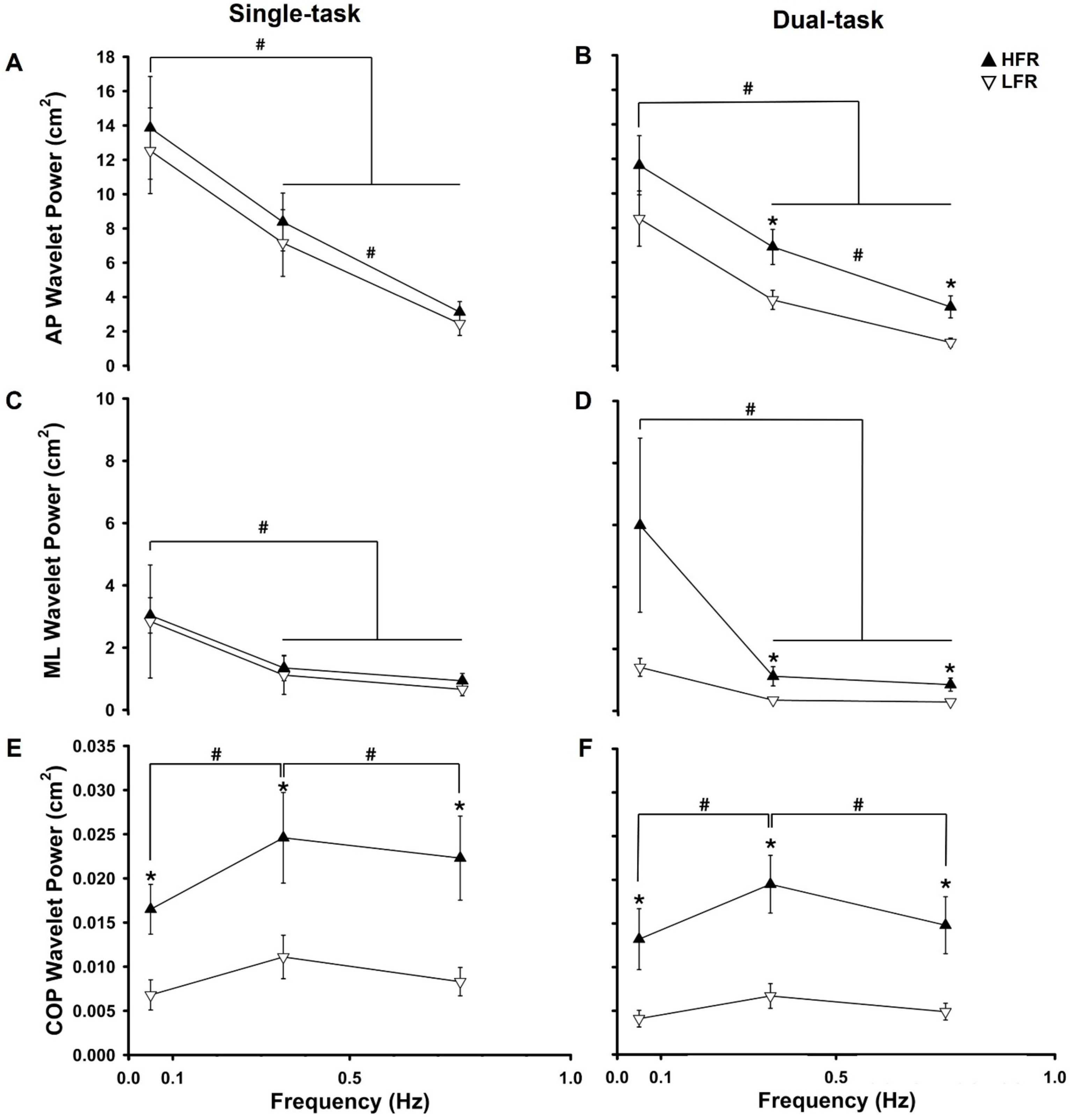
Absolute wavelet power spectrum of the COP 0-1 Hz signal. **(A-B)** AP wavelet power was significantly greater from 0-0.1 Hz when compared with 0.1-0.5 and 0.5-1 Hz, irrespective of task conditions. **(C-D)** ML wavelet power was significantly greater from 0-0.1 Hz when compared with 0.1-0.5 and 0.5-1 Hz, irrespective of task conditions. **(E-F)** Total COP wavelet power was significantly greater from 0.1-0.5 Hz when compared with 0-0.01 and 0.5-1 Hz, irrespective of task conditions. Older adults at HFR exhibited significantly greater total COP wavelet power when compared with LFR, irrespective of task conditions. # indicates a significant difference between frequency bands. * Indicates a significant difference between older adults at HFR and LFR

#### Medial-lateral 0-1 Hz: absolute and normalized wavelet power

In the absolute wavelet power for ML signal, the frequency main effect was significant (0-0.1 Hz: 3.32 ± 5.03 cm^2^, 0.1-0.5 Hz: 0.98 ± 1.29 cm^2^, 0.5-1 Hz: 0.682 ± 0.68 cm^2^; *F*_2,60_ = 9.3, *P* < 0.001, *η_p_*^2^= 0.237; Fig. 8C-D), indicating greater power from 0-0.1 Hz when compared with 0.1-0.5 and 0.5-1 Hz. In the normalized wavelet power spectrum, the frequency main effect was significant (0-0.1 Hz: 60.94 ± 11.02 %, 0.1-0.5 Hz: 19.55 ± 7.18 %, 0.5-1 Hz: 19.51 ± 9.33 %; *F*_2,60_ = 140.47, *P* < 0.001, *η_p_*^2^= 0.833), indicating greater power from 0-0.1 Hz when compared with 0.1-0.5 and 0.5-1 Hz. All other main effects and interactions were not significant.

#### Total COP 0-1 Hz: absolute and normalized wavelet power

In the absolute wavelet power for the total COP signal, the frequency main effect was significant (0-0.1 Hz: 0.01 ± 0.005 cm^2^, 0.1-0.5 Hz: 0.015 ± 0.012 cm^2^, 0.5-1 Hz: 0.013 ± 0.011 cm^2^; *F*_2,60_ = 5.4, *P* = 0.007, *η_p_*^2^= 0.153; Fig. 8E-F), indicating greater power from 0.1-0.5 Hz when compared with 0-0.1 and 0.5-1 Hz. The task main effect was significant (*F*_1,30_ = 4.5, *P* = 0.042, *η_p_*^2^= 0.131), indicating that both groups exhibited significantly greater COP absolute wavelet power under the single-task condition when compared with the dual-task condition. The group main effect was also significant (*F*_1,30_ = 17.3, *P* <.001, *η_p_*^2^= 0.366), demonstrating that the HFR group exhibited greater power across all frequency bands when compared with the LFR group, irrespective of task condition.

In the normalized wavelet power, the frequency main effect was significant (0-0.1 Hz: 26.92 ± 9.31 %, 0.1-0.5 Hz: 40.28 ± 7.36 %, 0.5-1 Hz: 32.80 ± 9.30 %; *F*_2,60_ = 12.6, *P* < 0.001, *η_p_*^2^= 0.296), indicating greater power from 0-1-0.5 Hz when compared with 0-0.1 and 0.5-1 Hz. All other main effects and interactions were not significant.

Association between absolute wavelet power spectrum from 0-1 Hz and total COP sway displacement.

We found that absolute wavelet power in the 0-1 Hz frequency band of the total COP sway displacement was significantly reduced during the dual-task condition when compared with the single-task condition. Furthermore, the HFR group exhibited significantly greater absolute and normalized wavelet power within this frequency band when compared with the LFR group. Therefore, we performed a multiple linear regression analysis to determine the association between absolute wavelet power in the 0-1 Hz band and total COP sway displacement. We found that the absolute wavelet power in the 0-1 Hz band is associated with an increase in total COP sway displacement irrespective of fall risk group (R^2^ = 0.58, *P* < 0.001; Fig. 9). To further investigate this relation, we performed an additional multiple linear regression analysis to determine the association between the absolute change in the wavelet power in the 0-1 Hz band from single to dual-task and total COP sway displacement. We found that the change in wavelet power in the 0-1 Hz band from single to dual-task is associated with an increase in total COP sway displacement (R^2^ = 0.57, *P* < 0.001; Fig. 10A). This relation indicates that modulation of the sway in the 0-1 Hz band accounts for ∼57% of the increase in sway from single to dual-task. Interestingly, this association was driven by the HFR group (R^2^ = 0.63, *P* < 0.001; Fig. 10B) but not by the LFR group (P > 0.7).

**Fig. 9.**
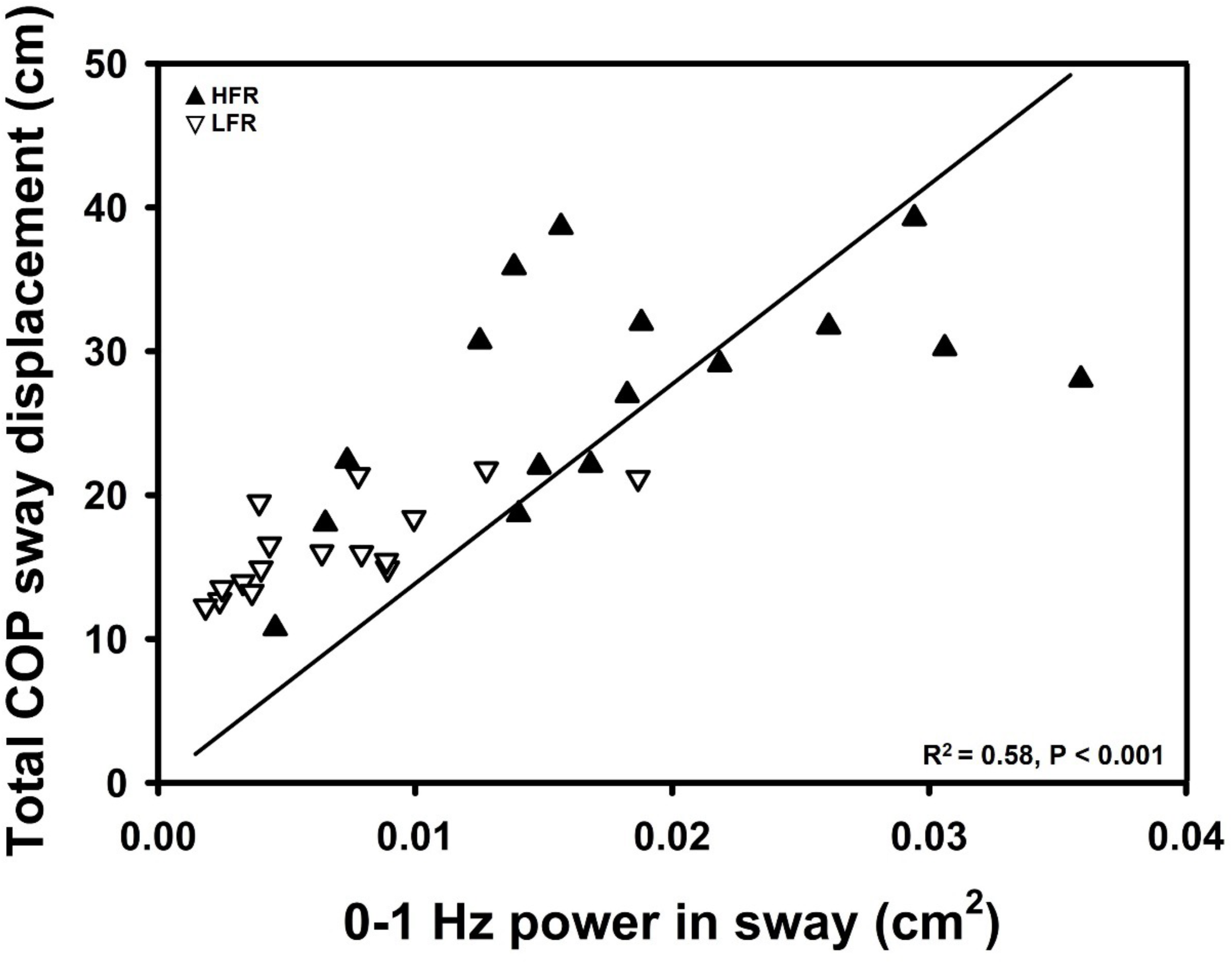
Association between the absolute wavelet power in the 0-1 Hz band and the total COP sway displacement. The absolute wavelet power in the 0-1 Hz band was associated with an increase in total COP sway displacement irrespective of fall risk group (R^2^ = 0.58, *P* < 0.001)

**Fig. 10.**
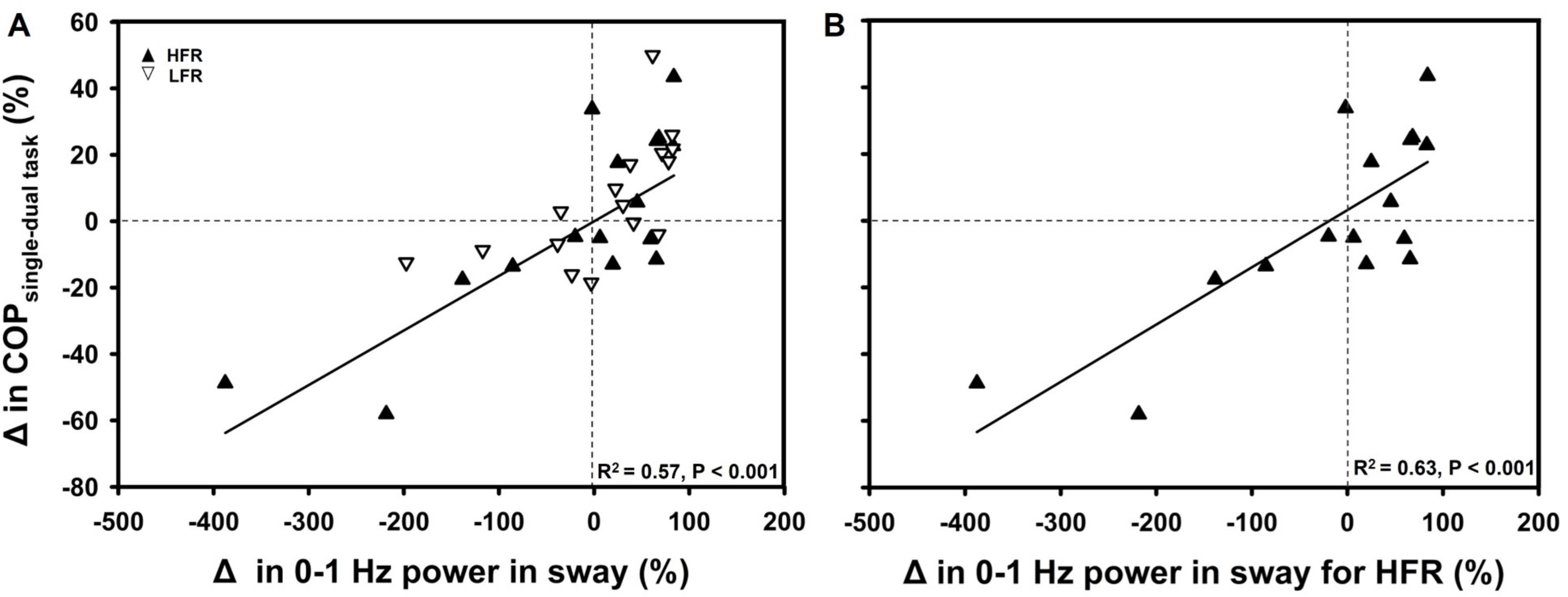
Association between the absolute change in wavelet power in the 0-1 Hz band from single to dual-task and total COP sway displacement. **(A)** The change in power in the 0-1 Hz band from single to dual-task is associated with an increase in total COP sway displacement (R^2^ = 0.57, *P* < 0.001). **(B)** The association between the change in power in the 0-1 Hz band from single to dual-task and the total COP sway displacement was driven by the HFR group (R^2^ = 0.63, *P* < 0.001).

## Discussion

The purpose of this study was to investigate the effects of cognitive dual-tasking on low-frequency oscillations during quiet standing in older adults. We explored oscillations from 0-4 Hz and 0-1 Hz frequency bands as potential predictors of fall risk in older adults to enhance our understanding of age-associated changes in postural control. It is well established that postural control declines with aging and that engaging in a dual-task could significantly increase postural sway, consequently elevating the risk of falls in older adults [1, 2, 44]. Therefore, we used this approach to determine whether dual-tasking disproportionately challenges postural sway in older adults at high risk for falls or if it universally challenges the aging population [5, 18, 45]. We found that: A) older adults at high fall risk exhibit greater postural sway when compared with older adults at low fall risk. B) Most of the absolute and normalized wavelet power from 0-4 Hz is concentrated within the 0-1 Hz frequency band across all directions. Specifically, ∼96% of the power in the anterior-posterior sway, ∼95% in medial-lateral sway, and ∼69% of the power in total COP sway come from the 0-1 Hz frequency range. C) The absolute change in wavelet power in the 0-1 Hz band from single to dual-task is associated with an increase in total COP sway displacement irrespective of fall risk group.

### Age-associated changes in postural sway and their relation to fall risk

We found that older adults at HFR exhibit greater postural sway in all directions (anterior-posterior, medial-lateral, and total COP sway displacement) when compared with the LFR group (Fig. 3). This supports and extends existing findings on age-associated changes in balance control mechanisms, including slower reaction times, decreased muscular strength, impaired sensory interaction, and increased susceptibility to fatigue – all factors that collectively contribute to greater postural instability [46, 47]. Furthermore, older adults at HFR exhibit significantly larger COP 95% ellipse area when compared with older adults at LFR (Fig. 3D). The increased ellipse area, which represents greater COP deviation, serves as a quantitative biomarker of postural instability [48]. However, it is essential to note that the postural sway metrics were not sensitive enough to differentiate between single and dual-task conditions. This limitation of linear measures sets the stage for our exploration of nonlinear measures in the subsequent analysis, where we found that bio-signatures in postural sway, particularly in low-frequency oscillations in the 0-1 Hz band, demonstrated sensitivity to both task conditions and age-associated changes.

### Age-associated changes in low-frequency oscillations and their relation to fall risk

Previous studies have found that most of the power in postural sway oscillations is concentrated from 0-4 Hz [17, 49]. However, other groups suggest that frequency oscillations in the 0-1 Hz band contain the most valuable information about the underlying mechanisms that constitute the COP signal [13, 19, 21]. To investigate this, we divided oscillations in postural sway into three frequency bands (0-1, 1-2, and 2-4 Hz). We found that older adults, irrespective of fall risk, exhibit significantly greater absolute and normalized wavelet power from 0-1 Hz (AP: ∼96 %, ML: ∼95 %, and COP: ∼69 %; Fig. 5-7) when compared with 1-2 Hz (AP: 3 %, ML: ∼4 %, and COP: ∼19 %), and 2-4 Hz (AP: ∼1 %, ML: ∼1 %, and COP: ∼12 %). Furthermore, we found that older adults at HFR exhibit significantly greater absolute and normalized wavelet power in total COP across all frequency bands when compared with the LFR group. Our findings partially align with those of Jafari et al. (2023), who indicated that older adults with sensorimotor decline and higher fall risk tend to rely on either very low-frequency or higher-frequency oscillations for postural control [13]. This demonstrates that modulation of postural sway differs with fall risk. However, contrary to this study, which concluded that older adults relying on low-frequency oscillations have greater postural control than those using higher-frequency oscillations, we found that older adults at HFR exhibited significantly greater power across all frequency bands. This increase in wavelet power across the frequency spectrum may represent a distinguishing characteristic of older adults at HFR, revealing limitations in compensatory strategies. Moreover, multiple linear regression analysis demonstrated that the absolute wavelet power in the 0-1 Hz band is associated with an increase in total COP sway displacement (R^2^ = 0.58, *P* < 0.001; Fig. 9). This finding corroborates previous research that has shown that the most critical information about postural sway is primarily contained in the 0-1 Hz frequency band [13, 19]. This underscores the crucial role of low-frequency oscillations in postural control mechanisms and supports their potential utility as biomarkers for fall risk assessments.

### Age-associated changes in postural sway oscillations during dual-task performance

We found no significant differences in the Hopkins Verbal Learning Test (HVLT) cognitive task between older adults at HFR when compared with those at LFR, whether sitting (single-task) or standing on the force plate (dual-task) (Fig. 4). This suggests that, irrespective of fall risk, both groups demonstrated comparable cognitive performance, indicating that the addition of the postural demand did not significantly impact cognitive task accuracy. However, when we analyzed postural sway oscillations at 0-1 Hz, we found that older adults, regardless of their fall risk status, exhibit significantly reduced COP wavelet power under the dual-task condition when compared with the single-task condition. Two factors could potentially explain these findings. Firstly, the cognitive task’s complexity: A five-word message without a non-meaningful connection may not provide sufficient cognitive load challenge to change postural control during the dual-task condition. This aligns with previous findings suggesting that non-meaningful auditory messages may not be challenging enough to modify postural control in older adults [50, 51].

Moreover, the relationship between cognition and balance is not exclusive to a single cognitive domain. Several groups have shown that executive function has the strongest association with balance, while memory is the weakest [52, 53]. Our task, which primarily engaged auditory processing and short-term memory, may not have sufficiently challenged the cognitive domains most relevant to postural control. Secondly, task prioritization: older adults may prioritize postural control over cognition performance, indicating an age-associated shift in functional priorities [54, 55]. These findings suggest that the relationship between dual-task and postural control is complex and context-dependent. It has been shown that, specifically in older adults, as postural threat increases, postural control temporarily improves while secondary task performance deteriorates [56]. This prioritization strategy paradoxically indicates an increased risk of falls. The reallocation of cognitive resources to maintain postural control, even during single-tasking, results in altered low-frequency oscillations. Consequently, when faced with complex real-world environments requiring divided attention, older adults may lack sufficient cognitive reserve to respond to unexpected balance challenges. Moreover, nonlinear measures, particularly low-frequency oscillations, proved to be more sensitive than linear postural sway metrics in detecting dual-task effects on the COP signal. Had we limited our analysis to linear postural sway measures, we would have incorrectly concluded that dual-tasking had no significant effect on postural control.

Moreover, this study provides evidence that the absolute change in wavelet power in the 0-1 Hz band, from single to dual-task, is associated with an increase in total COP sway displacement, irrespective of the fall risk group. This indicates that modulation of the sway at 0-1 Hz accounts for ∼57% of the increase in sway from single to dual-task (Fig. 10A), and this relation was driven only by the HFR group (Fig. 10B). This suggests that COP variability is primarily modulated by increased fall risk. In contrast, the LFR group did not show a significant association. Therefore, the model predominantly reflected the postural control mechanisms of individuals at HFR. These findings demonstrate that dual-tasking universally challenges postural control across the aging population, indicating that all older adults should exercise caution when performing a cognitive and balance task simultaneously. However, dual-tasking has a substantially greater impact on older adults at HFR.

In summary, our findings suggest that nonlinear postural sway measures provide valuable insights into age-associated changes in fall risk and dual-task performance. These findings have promising implications for clinical practice. Focusing on low-frequency oscillations, particularly in the 0-1 Hz range, could enable the earlier identification of individuals at high fall risk and a better understanding of how the dual-task paradigm challenges the aging population. This could enable timely interventions and the development of personalized fall prevention strategies. Future research should examine postural sway oscillations at 0 to 1 Hz employing more demanding cognitive tasks targeting executive functions to better understand the relation between dual-tasking and age-associated changes in postural control.

## Data Availability

All data produced in the present study are available upon reasonable request to the authors

## Acknowledgments

The authors have no acknowledgments to report.

## Author contributions

MMV – Data Curation, Formal analysis, Writing— Original Draft, Writing—Review & Editing, Visualization, HNR – Co-first author, Data Curation, Formal analysis, Writing— Original Draft, Writing—Review & Editing, Visualization, DJG - Conceptualization, Methodology, Writing—Original Draft, Writing—Review & Editing, PEG - Conceptualization, Methodology, Writing—Review & Editing, NV - Writing—Review & Editing, Supervision, NB-Writing—Review & Editing, Supervision, HSB - Conceptualization, Methodology, Resources, Data Curation, Formal analysis, Writing—Original Draft, Writing—Review & Editing, Supervision, Project administration, Funding acquisition.

## Declarations

### Ethics approval and consent to participate

All participants provided written informed consent before participating in the study. The ethics committee of San Diego State University approved this study.

## References

1. Goble DJ, Baweja HS. Normative Data for the BTrackS Balance Test of Postural Sway: Results from 16,357 Community-Dwelling Individuals Who Were 5 to 100 Years Old. Phys Ther. 2018; 98(9):779–785. 10.1093/ptj/pzy062.

2. Goble DJ, Baweja HS. Postural sway normative data across the adult lifespan: Results from 6280 individuals on the Balance Tracking System balance test. Geriatr Gerontol Int. 2018; 18(8):1225–1229. 10.1111/ggi.13452.

3. Vaishya R, Vaish A. Falls in Older Adults are Serious. Indian journal of orthopaedics. 2020; 54(1):69–74. 10.1007/s43465-019-00037-x.

4. Moreland B, Kakara R, Henry A. *Trends in Nonfatal Falls and Fall-Related Injuries Among Adults Aged ≥65 Years - United States*, *2012-2018*. MMWR. Morbidity and mortality weekly report. 2020; 69(27):875–881. 10.15585/mmwr.mm6927a5.

5. Chagdes JR, Rietdyk S, Haddad JM, Zelaznik HN, Raman A, Rhea CK, Silver TA. Multiple timescales in postural dynamics associated with vision and a secondary task are revealed by wavelet analysis. Exp Brain Res. 2009; 197(3):297–310. 10.1007/s00221-009-1915-1.

6. Pollind ML, Soangra R. Mini-Logger- A Wearable Inertial Measurement Unit (IMU) for Postural Sway Analysis. Annu Int Conf IEEE Eng Med Biol Soc. 2020:4600–4603. 10.1109/EMBC44109.2020.9175167.

7. Frames C, Soangra R, Lockhart TE. Assessment of postural stability using inertial measurement unit on inclined surfaces in healthy adults. Biomed Sci Instrum. 2013; 49:234–242.

8. Richmond SB, Dames KD, Goble DJ, Fling BW. Leveling the playing field: Evaluation of a portable instrument for quantifying balance performance. J Biomech. 2018; 75:102–107. 10.1016/j.jbiomech.2018.05.008.

9. O’Connor SM, Baweja HS, Goble DJ. Validating the BTrackS Balance Plate as a low cost alternative for the measurement of sway-induced center of pressure. J Biomech. 2016; 49(16):4142–4145. 10.1016/j.jbiomech.2016.10.020.

10. Pavol, MJ. Detecting and understanding differences in postural sway. Focus on “A new interpretation of spontaneous sway measures based on a simple model of human postural control”. J Neurophysiol. 2005; 93(1):20–21. 10.1152/jn.00864.2004.

11. Baltich J, Von Tscharner V, Zandiyeh P, Nigg BM. Quantification and reliability of center of pressure movement during balance tasks of varying difficulty. Gait & posture. 2014; 40(2):327–332. 10.1016/j.gaitpost.2014.04.208.

12. Collins JJ, De Luca CJ. Open-loop and closed-loop control of posture: A random-walk analysis of center-of-pressure trajectories. Exp Brain Res. 1993; 95:308–318. 10.1007/BF00229788.

13. Jafari H, Gustafsson T, Nyberg L, Röijezon U. Predicting balance impairments in older adults: a wavelet-based center of pressure classification approach. Biomed Eng Online. 2023; 22(1):83. 10.1186/s12938-023-01146-3.

14. Kyvelidou A, Harbourne RT, Shostrom VK, Stergiou N. Reliability of center of pressure measures for assessing the development of sitting postural control in infants with or at risk of cerebral palsy. Arch Phys Med Rehabil. 2010; 91(10):1593–1601. 10.1016/j.apmr.2010.06.027.

15. Assländer L, Peterka RJ. Sensory reweighting dynamics in human postural control. Journal of neurophysiology. 2014; 111(9):1852–1864. 10.1152/jn.00669.2013.

16. Pasma JH, Engelhart D, Maier AB, Schouten AC, van der Kooij H, Meskers CG. Changes in sensory reweighting of proprioceptive information during standing balance with age and disease. J Neurophysiol. 2015; 114(6):3220–3233. 10.1152/jn.00414.2015.

17. Hay DC, Wachowiak MP. Analysis of free moment and center of pressure frequency components during quiet standing using magnitude squared coherence. Hum Mov Sci. 2017; 54:101–109. 10.1016/j.humov.2017.04.002.

18. Kanekar N, Lee YJ, Aruin AS. Frequency analysis approach to study balance control in individuals with multiple sclerosis. J Neurosci Methods. 2014; 222:91–96. 10.1016/j.jneumeth.2013.10.020.

19. Redfern MS, Yardley L, Bronstein AM. Visual influences on balance. J Anxiety Disord. 2001; 15(1-2):81–94. 10.1016/s0887-6185(00)00043-8.

20. Yamamoto T, Smith CE, Suzuki Y, Kiyono K, Tanahashi T, Sakoda S, Morasso P, Nomura T. Universal and individual characteristics of postural sway during quiet standing in healthy young adults. Physiol Rep. 2015; 3(3):e12329. 10.14814/phy2.12329.

21. Kirchner M, Schubert P, Schmidtbleicher D, Haas CT. Evaluation of the temporal structure of postural sway fluctuations based on a comprehensive set of analysis tools. Physica A: Statistical Mechanics and its Applications. 2012; 391(20):4692–4703. 10.1016/j.physa.2012.05.034.

22. Peterka, RJ. Sensorimotor integration in human postural control. J Neurophysiol. 2002; 88(3):1097–1118. 10.1152/jn.2002.88.3.1097.

23. Stergiou N, Harbourne R, Cavanaugh J. Optimal movement variability: a new theoretical perspective for neurologic physical therapy. J Neurol Phys Ther. 2006; 30(3):120–129. 10.1097/01.npt.0000281949.48193.d9.

24. Salsabili H, Bahrpeyma F, Esteki A, Karimzadeh M, Ghomashchi H. Spectral characteristics of postural sway in diabetic neuropathy patients participating in balance training. J Diabetes Metab Disord. 2013; 12:29. 10.1186/2251-6581-12-29.

25. Goble DJ, Brown EC, Marks CR, Baweja HS. Expanded normative data for the balance tracking system modified clinical test of sensory integration and balance protocol. Medical Devices & Sensors. 2020; 3(3):e10084. 10.1002/mds3.10084.

26. Goble DJ, Brar H, Brown EC, Marks CR, Baweja HS. Normative data for the Balance Tracking System modified Clinical Test of Sensory Integration and Balance protocol. Med Devices (Auckl). 2019; 12 10.2147/MDER.S206530.

27. Wang J, Li Y, Yang GY, Jin K. Age-Related Dysfunction in Balance: A Comprehensive Review of Causes, Consequences, and Interventions. Aging and disease. 2024; 16(1):714–737. 10.14336/AD.2024.0124-1.

28. DL, Murman. The Impact of Age on Cognition. Seminars in hearing. 2015; 36(3):111–121. 10.1055/s-0035-1555115.

29. Brown LA, Shumway-Cook A, Woollacott MH. Attentional demands and postural recovery: the effects of aging. The journals of gerontology. Series A, Biological sciences and medical sciences. 1999; 54(4):165–171. 10.1093/gerona/54.4.m165.

30. Shumway-Cook A, Woollacott M. Attentional demands and postural control: the effect of sensory context. J Gerontol A Biol Sci Med Sci. 2000; 55(1):M10–M16. 10.1093/gerona/55.1.m10.

31. Prado JM, Stoffregen TA, Duarte M. Postural sway during dual tasks in young and elderly adults. Gerontology. 2007; 53(5):274–281. 10.1159/000102938.

32. Hadamus A, Błażkiewicz M, Kowalska AJ, Wydra KT, Grabowicz M, Łukowicz M, Białoszewski D, Marczyński W. Nonlinear and Linear Measures in the Differentiation of Postural Control in Patients after Total Hip or Knee Replacement and Healthy Controls. Diagnostics (Basel). 2022; 12(7):1595. 10.3390/diagnostics12071595.

33. Ghofrani M, Olyaei G, Talebian S, Bagheri H, Malmir K. Test-retest reliability of linear and nonlinear measures of postural stability during visual deprivation in healthy subjects. J Phys Ther Sci. 2017; 29(10) 10.1589/jpts.29.1766.

34. Turnock MJ, Layne CS. Variations in linear and nonlinear postural measurements under achilles tendon vibration and unstable support-surface conditions. J Mot Behav. 2010; 42(1):61–69. 10.1080/00222890903397103.

35. Yesavage JA, Brink TL, Rose TL, Lum O, Huang V, Adey M, Leirer VO. Development and validation of a geriatric depression screening scale: a preliminary report. J Psychiatr Res. 1982; 17(1):37–49. 10.1016/0022-3956(82)90033-4.

36. Goble DJ, Khan E, Baweja HS, O’Connor SM. A point of application study to determine the accuracy, precision and reliability of a low-cost balance plate for center of pressure measurement. J Biomech. 2018; 71:277–280. 10.1016/j.jbiomech.2018.01.040.

37. Goble DJ, Baweja N, Baweja HS. BTrackS: A Low-Cost, Portable Force Plate for Objectively Measuring Balance Deficits and Fall Risk. Home Healthc Now. 2019; 37(6):355–356. 10.1097/NHH.0000000000000823.

38. Brandt, J. The Hopkins Verbal Learning Test: Development of a new memory test with six equivalent forms. Clinical Neuropsychologist. 1991; 5(2):125–142. 10.1080/13854049108403297.

39. Dault MC, Yardley L, Frank JS. Does articulation contribute to modifications of postural control during dual-task paradigms? Brain Res Cogn Brain Res. 2003; 16(3):434–440. 10.1016/s0926-6410(03)00058-2.

40. Torrence C, Compo GP. A practical guide to wavelet analysis. Bulletin of the American Meteorological Society. 1998; 79:61–78.

41. Baweja HS, Patel BK, Neto OP, Christou EA. The interaction of respiration and visual feedback on the control of force and neural activation of the agonist muscle. Human movement science. 2011; 30(6):1022–1038. 10.1016/j.humov.2010.09.007.

42. Addison, PS, The Illustrated Wavelet Transform Handbook. 1st Edition ed. 2002, Boca Raton. 464.

43. Grinsted A, Moore JC, Jevrejeva S. Application of the cross wavelet transform and wavelet coherence to geophysical time series. Processes Geophys. 2004; 11(5/6):561–566. 10.5194/npg-11-561-2004.

44. Bergamin M, Gobbo S, Zanotto T, Sieverdes JZ, Alberton CL, Zaccaria M, Ermolao A. Influence of age on postural sway during different dual-task conditions. Frontiers in aging neuroscience. 2014; 6:271. 10.3389/fnagi.2014.00271.

45. Cabeza-Ruiz R, García-Massó X, Centeno-Prada RA, Beas-Jiménez JD, Colado JC, González LM. Time and frequency analysis of the static balance in young adults with Down syndrome. Gait & Posture. 2011; 33(1):23–28. 10.1016/j.gaitpost.2010.09.014.

46. Morrison S, Colberg SR, Parson HK, Neumann S, Handel R, Vinik EJ, Paulson J, Vinik AI. Walking-Induced Fatigue Leads to Increased Falls Risk in Older Adults. J Am Med Dir Assoc. 2016; 17(5):402–409. 10.1016/j.jamda.2015.12.013.

47. Zhang S, Xu W, Zhu Y, Tian E, Kong W. Impaired Multisensory Integration Predisposes the Elderly People to Fall: A Systematic Review. Front Neurosci. 2020; 14:411. 10.3389/fnins.2020.00411.

48. Sargent C, Darwent D, Ferguson SA, Roach GD. Can a simple balance task be used to assess fitness for duty? Accid Anal Prev. 2012; 45:74–79. 10.1016/j.aap.2011.09.030.

49. Soames RW, Atha J. The spectral characteristics of postural sway behaviour. Eur J Appl Physiol Occup Physiol. 1982; 49(2) 10.1007/BF02334065.

50. Deviterne D, Gauchard GC, Jamet M, Vançon G, Perrin PP. Added cognitive load through rotary auditory stimulation can improve the quality of postural control in the elderly. Brain Res Bull. 2005; 64(6):487–492. 10.1016/j.brainresbull.2004.10.007.

51. Huxhold O, Li SC, Schmiedek F, Lindenberger U. Dual-tasking postural control: aging and the effects of cognitive demand in conjunction with focus of attention. Brain Res Bull. 2006; 69(3):294–305. 10.1016/j.brainresbull.2006.01.002.

52. Divandari N, Bird ML, Vakili M, Jaberzadeh S. The Association Between Cognitive Domains and Postural Balance among Healthy Older Adults: A Systematic Review of Literature and Meta-Analysis. Curr Neurol Neurosci Rep. 2023; 11(6):681–693. 10.1007/s11910-023-01305-y.

53. Sullivan EV, Rose J, Rohlfing T, Pfefferbaum A. Postural sway reduction in aging men and women: relation to brain structure, cognitive status, and stabilizing factors. Neurobiol Aging. 2009; 30(5):793–807. 10.1016/j.neurobiolaging.2007.08.021.

54. Zawadka-Kunikowska M, Klawe JJ, Tafil-Klawe M, Bejtka M, Rzepiński Ł, Cieślicka M. Cognitive Function and Postural Control Strategies in Relation to Disease Progression in Patients with Parkinson’s Disease. Int J Environ Res Public Health. 2022; 19(19):12694. 10.3390/ijerph191912694.

55. Tsang WW, Chan VW, Wong HH, Yip TW, Lu X. The effect of performing a dual-task on postural control and selective attention of older adults when stepping backward. J Phys Ther Sci. 2016; 28(10):2806–2811. 10.1589/jpts.28.2806.

56. Brown LA, Sleik RJ, Polych MA, Gage WH. Is the prioritization of postural control altered in conditions of postural threat in younger and older adults?. The journals of gerontology. Series A, Biological sciences and medical sciences. 2002; 57(12):M785–M792. 10.1093/gerona/57.12.m785.

